# Wildfire exposure and academic performance in Brazil: a causal inference approach for spatiotemporal data

**DOI:** 10.1101/2023.05.09.23289704

**Authors:** Sean McGrath, Rajarshi Mukherjee, Weeberb J. Réquia, Wan-Chen Lee

**Affiliations:** Department of Biostatistics, Harvard T.H. Chan School of Public Health, Boston, USA; School of Public Policy and Government, Fundação Getúlio Vargas, Brasília, Brazil; Institute of Environmental and Occupational Health Sciences, College of Public Health, National Taiwan University, Taipei, Taiwan

**Keywords:** wildfire, adolescents, academic performance, causal inference, spatiotemporal data

## Abstract

As the frequency and intensity of wildfires are projected to globally amplify due to climate change, there is a growing need to quantify the impact of exposure to wildfires in vulnerable populations such as adolescents. In our study, we applied rigorous causal inference methods to estimate the effect of wildfire exposure on academic performance of high school students in Brazil between 2009 and 2015. Using longitudinal data from 8,183 high schools across 1,571 municipalities in Brazil, we estimated that the average performance in most academic subjects decreases under interventions that increase wildfire exposure, e.g., a decrease of 1.8% (*p* = 0.01) in the natural sciences when increasing the wildfire density from 0.0035 wildfires/km^2^ (first quantile in the sample) to 0.0222 wildfires/km^2^ (third quartile). Furthermore, these effects considerably worsened over time and were worse in regions with lower income. Our findings highlight the adverse impact of wildfires on educational outcomes.

## 1. INTRODUCTION

Exposure to air pollution source emissions has been linked with both acute (Lee et al. 2016; Weichenthal, Hatzopoulou, and Goldberg 2014) and chronic (Laumbach and Kipen 2012; Xu et al. 2016) health effects. Vulnerable population groups such as people with pre-existing health conditions (Pope et al. 2015), older adults (Bell, Zanobetti, and Dominici 2014), and children (Du et al. 2012; Svendsen et al. 2012) are suggested to be at higher risk for health effects compared to the general population.

Air quality surrounding schools is a crucial concern worldwide. A growing body of evidence has reported that numerous schools are located in regions with high levels of air pollutants, posing a risk to children’s health (Guo et al. 2010; Richmond-Bryant et al. 2009; Rivas et al. 2014). The adverse effects of poor air quality on children’s health are not limited to general health outcomes such as respiratory and circulatory diseases (Andersen et al. 2007; Fan et al. 2016); studies have also linked it to cognitive outcomes in school-aged children (Calderon-Garciduenas et al. 2008; Martinez-Lazcano et al. 2013; Suglia et al. 2007). Cognitive performance, such as cognitive development and working memory, is the primary indicator of these outcomes (Forns et al. 2017; van Kempen et al. 2012). Some epidemiological investigations have even used academic achievement as a proxy for cognitive performance to estimate the impact of poor air quality on students (Mohai et al. 2011).

Wildfire is a source of air pollution that involves many potentially hazardous conditions for children. Wildfires emit or result in a mix of primary and secondary pollutants-particulate pollution (including PM_2.5_ and its elemental and black carbon components), gases (e.g., CO, NO_2_, O_3_), and, depending on what products are burned, there are emissions of other toxic pollutants like benzene or formaldehyde (Balmes 2018). Evidence has shown that each air pollutant has different toxicologic and physiologic effects on human health (Brook et al. 2010). Emissions from forest fires can travel over large distances, affecting air quality and human health far from the originating fires (Youssouf et al. 2014). The frequency and intensity of wildfires were projected to be amplified by climate and land-use changes, resulting in a global increase of extreme fires of up to 14% by 2030 and 50% by the end of the century (Sullivan, Baker, and Kurvits 2022). The mutually exacerbating relationship between wildfires and climate change highlights the urgent need for understanding the impacts of wildfire exposure and its associated public health outcomes.

Previous studies have investigated the relationship between exposure to wildfires and students’ academic achievements. Some of these studies performed detailed association analyses and did not attempt to estimate causal effects (e.g., see Reid et al. (2016)), while others such as Wen and Burke (2022), Cleland et al. (2022), Miller and Hui (2022), and Gibbs et al. (2019) aimed to estimate causal effects by applying traditional causal inference methods developed for independent and identically distributed data (Hernán and Robins 2010). Several major challenges arise when applying traditional causal inference methods in this setting due to the spatiotemporal nature of the data, which we elaborate on in Section 2.2. In brief, such analyses in spatiotemporal settings have imprecise definitions of causal effects and require researchers to make strong and often untenable assumptions on the causal structure and distribution of the data.

To address this gap, in this paper, we apply rigorous causal inference methods to estimate the average causal effect of exposure to wildfires on academic performance of high school students in Brazil. Specifically, we adopt the causal inference framework for spatiotemporal data recently developed by Christiansen et al. (2022). By doing so, our analyses make considerably weaker causal assumptions and distributional assumptions compared to those from other frameworks, and our analyses estimate well-defined causal effects. We first apply this framework to estimate the impact of wildfire exposure on performance in five different academic subjects in Brazil’s High School National Exam between 2009 and 2015. Then, we quantify how these effects change over time and change across different geographic regions of Brazil, defined by their average income and exposure to wildfires.

## 2. MATERIALS AND METHODS

### 2.1. Data

#### 2.1.1. School-level academic performance

First, we assessed the list of addresses (including street number, city, state, and postal code) of all schools in Brazil (more than 180 thousand schools). This list was obtained from the National Institute for Educational Research in Brazil, known as INEP - *Instituto Nacional de Estudos e Pesquisas Educacionais Anísio Teixeira*. This institute is a governmental agency under the Brazilian Ministry of Education. Then, using the addresses from each school, we geocoded the data by spatially creating the latitude and longitude for each school and displaying the schools over Brazil.

Then, the information on academic performance at the school level was linked to the corresponding schools geocoded in the first stage. School-level academic performance was obtained from the High School National Exam, shortened as ENEM in Brazil (*Exame Nacional do Ensino Medio*). The ENEM is an annual (held between November and December), non-mandatory, and standardized Brazilian national exam managed by the INEP to evaluate the level of knowledge of high school students in Brazil. The ENEM exam includes more than 150 multiple-choice questions in the following subjects: natural sciences (chemistry, physics, and biology), human sciences (geography, history, philosophy, sociology, and general knowledge), mathematics, and Portuguese language (grammar, reading comprehension, and literature). Also, the students must write an essay. The ENEM is used as an admission exam for enrollment in several universities in Brazil. On average, about 3.5 million students take the ENEM test annually.

The ENEM data provided by the INEP includes the annual average of ENEM scores by schools for each subject between 2009 and 2015. We used these averages to represent the school-level academic performance (SAP) in our analysis. The SAP ranges from 0 to 1,000, of which 1,000 indicates the highest ENEM score. The INEP calculated these averages only for high schools that had at least 2% of students taking the ENEM test. After applying this inclusion criterion, the final dataset encompassed 10,518 schools across Brazil. Figure 1 shows the spatial distribution of these schools.

**Figure 1.**
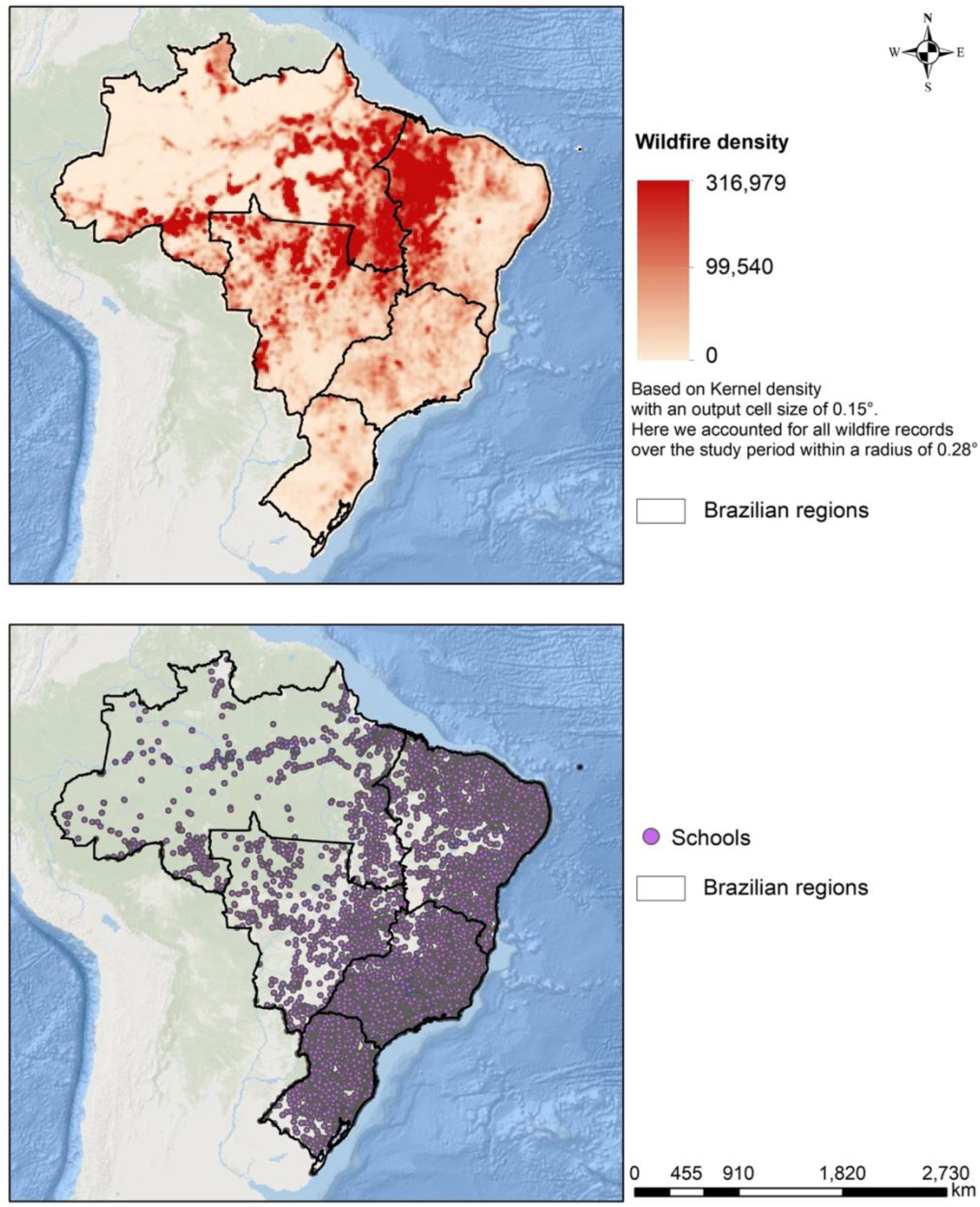
Spatial distribution of wildfire density and schools across Brazil.

#### 2.1.2. Wildfire

Wildfire data were provided by the National Institute of Spatial Research of Brazil – *Instituto Nacional de Pesquisas Espaciais - INPE* (http://queimadas.dgi.inpe.br/queimadas/). The data include the date of wildfire records and its geographical location. These data are derived from seven satellite remote sensing observations, including NOAA-18, NOAA-19, METOP-B, MODIS (NASA TERRA and AQUA), VIIRS (NPP-Suomi and NOAA-20), GOES-16, and MSG-3. All images from these satellites are processed by INPE to estimate wildfire records. This estimate is based on a specific satellite as a reference – the AQUA satellite. We accounted for all wildfire records in Brazil based on the reference satellite in the period between 2009 and 2015. Then, we used GIS techniques to summarize the annual number of wildfire occurrences within each municipality in Brazil. Finally, we estimated the spatial density of wildfires within each municipality by dividing the total number of wildfires in the municipality by the total area of the municipality.

### 2.2 Causal inference for spatiotemporal data

Since our study aims to estimate causal effects of wildfire exposure on academic performance (rather than just their associations), we need to operate under a suitable causal inference framework. Approaches for estimating causal effects require adjusting for confounders of the relationship between wildfire exposure and academic performance, such as socioeconomic status, prior academic achievement, health status, school quality, and environmental/climate conditions. The relationship between wildfire exposure and academic performance is also confounded by spatial factors, such as the location of the schools and students’ homes, which can affect both the students’ exposure to wildfires and their academic performance, as well as the availability of resources and social support systems that may mitigate the negative effects of wildfire exposure. Not accounting for such geographical and spatial factors would introduce bias in estimates of the causal effect of wildfire exposure on academic performance.

Traditional causal inference studies often operate under frameworks that were developed for independent and identically distributed data (e.g., structural causal models, causal graphical models, and the potential outcomes framework) (Hernán and Robins 2010). These frameworks are challenging to apply for spatiotemporal data, as there are usually complex dependencies in spatiotemporal data. For example, these frameworks rely on researchers to specify the full causal structure of the data, which is especially challenging in settings with spatiotemporal data. There may be unobserved spatial confounders of the relationship between the exposure and outcome, which would violate the assumption of knowing the full causal structure of the data. Moreover, applications of these frameworks in settings with spatiotemporal data often make strong distributional assumptions and do not have precise definitions of causal effects.

For estimating causal effects based on spatiotemporal exposure-outcome data, most recent research has focused on extending the concept of Granger causality (Granger 1980; Wiener 1956) to spatiotemporal data settings. In these methods, one reduces questions about causal effects to prediction type questions Moreover, most existing methods do not operate under a formal model for the causal structural of the spatiotemporal data and consequently do not have a precise and interpretable definition of a causal effect. To address these shortcomings, we adopt a recent causal inference framework for spatiotemporal developed by Christiansen et al. (2022) which formally defines causal effects while making very weak assumptions compared to prior attempts. This framework does not make any particular distributional assumptions on the data and accommodates autocorrelations in the outcome variable. Moreover, this framework does not require fully specifying the causal structure of the data. For example, this framework accounts for any (even if unmeasured) confounders of the relationship between the exposure and outcome provided that the confounders do not vary over time, such as genetic factors, certain demographic variables such as sex and race, and fixed environmental characteristics such as altitude or distance to a coastline. Furthermore, they develop a valid estimator of average causal effects and a corresponding hypothesis test under this framework, which we describe in more detail in the following subsection.

### 2.3. Statistical analysis

#### 2.3.1 Estimating the impact of wildfire exposure on academic performance

We applied the approach of Christiansen et al. (2022) to estimate the Average Causal Effect (ACE) of exposure to wildfire on academic performance in each of the five subjects (natural sciences, human sciences, mathematics, Portuguese language, and essay writing). This approach can be described in our setting as follows.

First, we fit a linear regression model separately at each municipality, where the outcome is SAP for a given academic subject and the predictor is wildfire density. For academic subject *z* and municipality *m*, the model is represented by

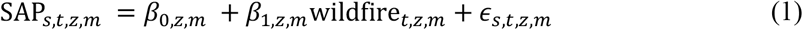

where SAP_*s,t,z,m*_ is the SAP for school *s* in municipality *m* measured at the end of year *t* (2009-2015), wildfire_*t,z,m*_ is the wildfire density in municipality *m* during year *t*, and *ϵ*_*s,t,z,m*_ is a mean-zero random error for school *s* in municipality *m* during year *t*. The regression coefficients (*β*_0,*z,m*_, *β*_1,*z,m*_) do not have causal interpretations and are eventually used to compute the ACE estimates. We truncated the modelled mean of SAP to fall within the range from 0 to 1000, as SAP values beyond this range are not feasible. Note that this approach adjusts for all possible time-invariant confounders of the relationship between academic performance and wildfire exposure because a different regression model is fit in each municipality. We estimated the average academic performance under any intervention that sets the wildfire density of all municipalities to level *w* by taking the average of the municipality-specific modelled means for SAP at wildfire density *w*.

For each academic subject, we estimated the ACE comparing wildfire exposure levels *w* = 0.0035 wildfires/km^2^ (which we refer to as *low density of wildfires*) and *w* = 0.0222 wildfires/km^2^ (which we refer to as *high density of wildfires*). These values were chosen by taking the first and third sample quartiles, respectively, of the municipality-specific wildfire densities. The ACE can be interpreted as the average academic performance (in the given subject) that would have been observed under any intervention that sets the wildfire density of all municipalities to the high level minus the average academic performance that would have been observed under any intervention that sets the wildfire density to the low level.

We applied the nonparametric resampling test of Christiansen et al. (2022) to test the null hypothesis of no ACE for each of the academic subjects. This approach involves creating *B* pseudo-replications of the observed data set by permuting the SAP values within each municipality. A p-value for the test is obtained by comparing the ACE estimate obtained from the observed data set to the ACE estimates obtained from the pseudo-replications. We used 250 pseudo-replications for computational reasons.

#### 2.3.2 Subgroup analyses: Estimating how the impact of wildfire exposure changes over time and space

We performed subgroup analyses to evaluate how the effects of wildfire exposure on academic performance may vary over time and space. Specifically, we performed the analyses described in the previous subsection to estimate ACEs based on data prior to (and including) a given cutoff year versus data after the cutoff year. The following cutoff years were considered: 2010, 2011, 2012, and 2013. Moreover, we performed subgroup analyses based on the municipalities’ average wildfire exposure and average income. Specifically, we estimated ACEs among municipalities with an average wildfire density below the median level (0.0125 wildfires/km^2^) and estimated ACEs among municipalities with an average wildfire density above the median level. Similarly, we estimated ACEs among municipalities with an average income below (and above) the median level (i.e., R$797.71, currency of Brazil).

#### 2.3.3 Sensitivity analyses

We performed several sensitivity analyses to explore the robustness of our estimates. First, we used different ranges for the modelled mean SAP values in the municipality-specific regression models: (i) 0 to 800, (ii) 200 to 1000, and (iii) 200 to 800. Second, we performed a log transformation of the wildfire density in the municipality-specific regression models. Third, we estimated the ACEs by taking the sample median (rather than the sample mean) of the municipality-specific estimates. These sensitivity analyses aim to help mitigate the impact of extrapolation beyond the observed range of the wildfire density values in the municipality-specific regression models when estimating the ACEs.

These analyses were performed in R (version 4.2.2).

## 3. RESULTS

### 3.1 Descriptive analyses

Wildfire exposure and academic performance data were available on 10,064 high schools in 3,110 municipalities in Brazil. We excluded data from any municipalities (1,539 municipalities) that had data available for less than two distinct years, as at least two years are required to fit the municipality-specific regression model. Data from 8,183 high schools across 1,571 municipalities in Brazil were included in our analyses.

A total of 5,046 (62%) of these schools were public (versus private) and 7,858 (96%) were located in urban areas (versus rural areas). Additionally, 1,940 (24%) of these schools had less than 30 students, 1,934 (24%) had between 31 and 60 students, 1,208 (15%) had between 61 and 90 students, and 3,101 (38%) had more than 90 students.

Figure 1 illustrates the spatial distribution of wildfire density and schools across Brazil. Figure 2 shows the average wildfire density and the average performance in each academic subject during each year of our study. While there was a moderate increase in the average wildfire density in 2010, there were generally no clear time trends for these variables.

**Figure 2.**
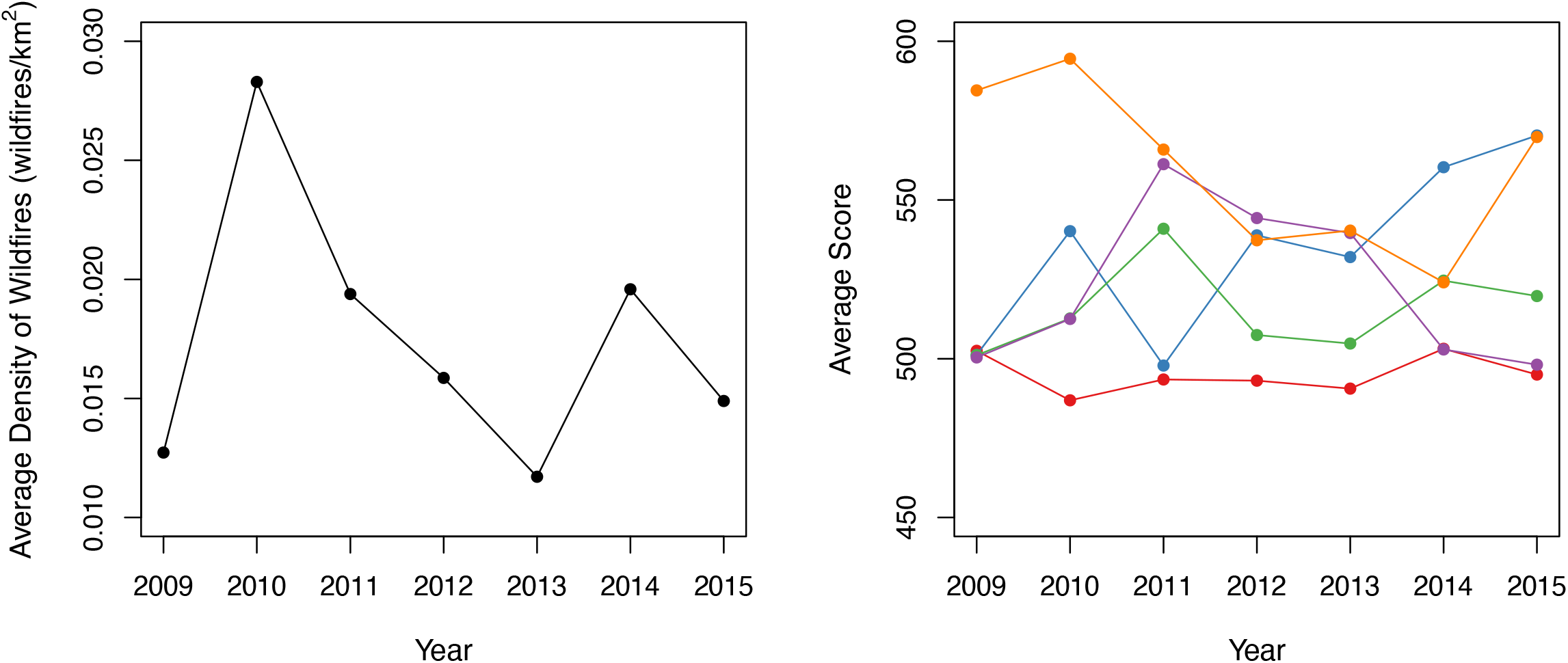
Time trends in the average density of wildfires (left panel) and academic performance (right panel). In the right panel, the red line corresponds to natural sciences scores; the blue line corresponds to human sciences scores; the green line corresponds to Portuguese language scores; the purple line corresponds to mathematics scores; the orange line corresponds to essay scores.

### 3.2: Impact of wildfire exposure on academic performance

In most academic subjects, the estimated average academic performance was higher under the intervention that sets wildfire exposure to the low level compared to the intervention that sets wildfire exposure to the high level (Figure 3). Specifically, the average academic performance increased by 4.23 points (0.88%, 479.48 points vs 483.71 points, *p* = 0.06) in natural sciences, 9.43 points (1.83%, 514.95 points vs 524.38 points, *p* = 0.01) in social sciences, 3.31 points (0.67%, 495.05 points vs 498.36 points, *p* = 0.18) in Portuguese language, and 6.16 points (1.25%, 494.47 points vs 500.63 points, *p* = 0.10) in mathematics under the intervention setting wildfire exposure to the low level. However, the estimated average academic performance decreased by 14.05 points (2.57%, 545.95 points vs 531.89 points, *p* = 0.01) in essay writing.

**Figure 3.**
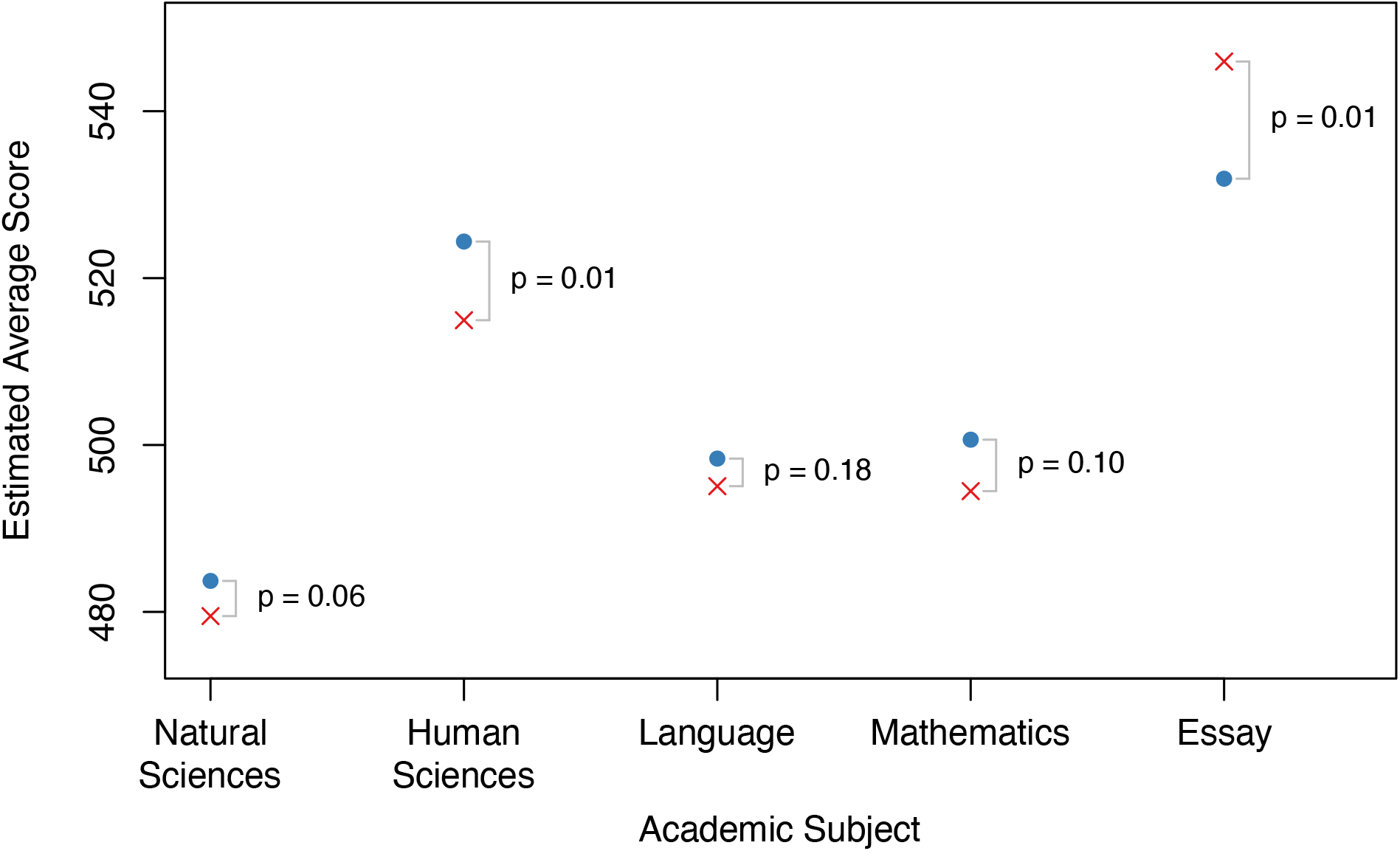
Estimated average score in each academic subject under the intervention setting the wildfire density of all municipalities to the low level (blue dots) and the high level (red x marks). The p-values correspond to the nonparametric permutation test of the null hypothesis of no average causal effect comparing the low and high levels of wildfire density.

### 3.3: How the impact of wildfire exposure changes over time and space

The results of the subgroup analyses are presented in Table 1. The ACE estimates typically decreased over time, indicating that the impact of wildfire exposure on academic performance became more harmful over time. For instance, in when comparing data prior to and including 2012 versus after 2012, the ACE estimates decreased by 13.70 points (*p* = 0.04) in natural sciences, 28.03 points (*p* = 0.01) in human sciences, 18.77 points (*p* = 0.02) in Portuguese language, 19.07 points (*p* = 0.02) in mathematics, and 36.83 points (*p* = 0.01) in essay writing. Similar trends were observed for other cutoff years.

**Table 1.** Estimates of the average causal effect (ACE) comparing low and high levels of wildfire exposure on academic performance.

| Subgroup | ACE Estimate (p-value) |  |  |  |  |
| --- | --- | --- | --- | --- | --- |
|  | Natural Sciences | Human Sciences | Language | Mathematics | Essay |
| All municipalities | -4.23 (0.06) | -9.43 (0.01) | -3.31 (0.18) | -6.16 (0.10) | 14.05 (0.01) |
| By year (2010) |  |  |  |  |  |
| Pre 2010<sup>1</sup> | 3.24 (0.54) | 14.82 (0.02) | 6.53 (0.18) | 12.11 (0.02) | 10.43 (0.05) |
| Post 2010 | -2.72 (0.37) | -14.49 (0.02) | 3.98 (0.15) | -5.25 (0.29) | -10.65 (0.10) |
| Change | -5.96 (0.33) | -29.31 (0.01) | -2.55 (0.53) | -17.36 (0.04) | -21.08 (0.01) |
| By year (2011) |  |  |  |  |  |
| Pre 2011 | 0.72 (0.96) | 24.70 (0.01) | 8.23 (0.07) | 6.57 (0.20) | 17.28 (0.01) |
| Post 2011 | -0.52 (0.84) | -2.05 (0.81) | -3.22 (0.47) | -15.51 (0.02) | -14.92 (0.01) |
| Change | -1.24 (0.87) | -26.75 (0.01) | -11.45 (0.02) | -22.08 (0.01) | -32.19 (0.01) |
| By year (2012) |  |  |  |  |  |
| Pre 2012 | 1.37 (0.78) | 10.95 (0.02) | 6.57 (0.08) | -0.82 (0.99) | 17.45 (0.01) |
| Post 2012 | -12.33 (0.03) | -17.08 (0.02) | -12.20 (0.04) | -19.89 (0.01) | -19.38 (0.02) |
| Change | -13.70 (0.04) | -28.03 (0.01) | -18.77 (0.02) | -19.07 (0.02) | -36.83 (0.01) |
| By year (2013) |  |  |  |  |  |
| Pre 2013 | 4.26 (0.26) | 6.00 (0.10) | 8.93 (0.01) | -5.35 (0.21) | 24.72 (0.01) |
| Post 2013 | 1.24 (0.89) | -14.44 (0.03) | -0.64 (0.97) | -0.86 (0.98) | -14.34 (0.28) |
| Change | -3.03 (0.80) | -20.44 (0.02) | -9.57 (0.19) | 4.49 (0.61) | -39.06 (0.01) |
| By average exposure level<sup>2</sup> |  |  |  |  |  |
| Below the median | -5.28 (0.35) | -13.53 (0.06) | -6.43 (0.14) | -7.12 (0.33) | 18.63 (0.01) |
| Above the median | -3.24 (0.10) | -5.56 (0.03) | -0.36 (0.80) | -5.26 (0.04) | 9.73 (0.02) |
| Change | 2.04 (0.84) | 7.97 (0.35) | 6.07 (0.23) | 1.86 (0.94) | -8.91 (0.14) |
| By average income level<sup>3</sup> |  |  |  |  |  |
| Below the median | -4.69 (0.12) | -8.77 (0.02) | -4.93 (0.13) | -9.77 (0.01) | 13.59 (0.01) |
| Above the median | -3.44 (0.59) | -13.23 (0.02) | 1.37 (0.67) | 6.08 (0.27) | 14.79 (0.01) |
| Change | 1.25 (0.75) | -4.46 (0.64) | 6.31 (0.25) | 15.85 (0.04) | 1.19 (0.80) |

<sup>1</sup> The subgroup “Pre 2010” includes all data before and including 2010. The subgroups “Pre 2011”, “Pre 2012”, and “Pre 2013” were defined in the same manner.
<sup>2</sup> The median wildfire density was 0.0125 wildfires/km<sup>2</sup>.
<sup>3</sup> The median income level was R\$797.71 (currency of Brazil).

In the subgroup analyses comparing the ACE estimates between municipalities with different levels of wildfire exposure (i.e., above or below the median wildfire density), the ACE estimates were further from 0 in municipalities with an average wildfire density below the median, although these differences were often not large. Compared to municipalities with an average wildfire density above the median, the ACE estimates in municipalities with an average wildfire density below the median exposure decreased by 2.04 points (*p* = 0.84) in natural sciences, 7.97 points (*p* = 0.35) in human sciences, 6.07 points (*p* = 0.23) in Portuguese language, and 1.86 points (*p* = 0.94) in mathematics; the ACE estimate increased by 8.91 points (*p* = 0.14) in essay writing in municipalities with an average wildfire density below the median.

In the subgroup analyses based on the municipalities’ average income, the ACE estimates in most academic subjects were more negative (i.e., the impact of wildfire exposure is more harmful) for municipalities with an average income below the median compared to those with an average income above the median. The ACE estimates for municipalities with lower income were 1.25 points lower in natural sciences (*p* = 0.75), 4.46 points higher in human sciences (*p* = 0.64), 6.31 points lower in Portuguese language (*p* = 0.25), 15.85 points lower in mathematics (*p* = 0.04), and 1.19 points lower in essay writing (*p* = 0.80).

### 3.4 Sensitivity analyses

The results of the sensitivity analyses are given in Supplementary Tables 1-5. The ACE estimates for all subjects typically decreased when using an upper bound of 800 for the modelled mean SAP, and they typically increased when using a lower bound of 200 for the modelled mean SAP. The ACE estimates were not strongly affected when performing a log transformation of the wildfire density in the municipality-specific regression models. When estimating the ACEs by taking the median of the municipality-specific estimates, the ACE estimates were often closer to 0.

## 4. DISCUSSION

Our study applied rigorous causal inference methods to estimate the impact of exposure to wildfires on the academic performance of high school students in Brazil. The results of our analyses indicate that increased exposure to wildfires worsens students’ performance in most academic subjects. Moreover, we found that such effects are considerably worsening over time and are worse in regions with lower income.

Although there is limited research on the association between wildfires and academic performance, a systematic review suggested that exposure to traffic-related air pollutants could affect students’ academic performance (Stenson et al. 2021). To explain the impact of traffic-related air pollution on academic performance, Stenson et al. (2021) proposed a conceptual framework that includes a direct pathway through impact on cognitive function and an indirect pathway involving increased school absences due to ill-health, such as worsening of asthmatic symptoms. The impact of wildfire and traffic emissions on academic performance may share some comparable pathways, as both arise from combustion processes, albeit with different compositions and toxicity of particulate matter components (Verma et al. 2009; Aguilera et al. 2021; Adetona et al. 2016; Wu, Jin, and Carlsten 2018). Increased respiratory admission among the elderly population has been linked to wildfire PM_2.5_ exposure, as the concentration of PM_2.5_ tends to be higher during wildfire episodes (Liu et al. 2017). However, more recent research has shown that even at similar exposure levels, PM_2.5_ from wildfires poses a significantly greater risk to respiratory health compared to other ambient sources for the general population. The authors further concluded that PM_2.5_ originating from wildfires may pose significant risks up to 10 times higher than PM_2.5_ from other ambient sources (Aguilera et al. 2021).

The subgroup analysis conducted in our study revealed that students from lower income families were more vulnerable to the wildfire impacts. A possible explanation for this finding is that income is closely related to the quality of housing, including the permeability of the building envelope and access to mechanical ventilation (Chan, Joh, and Sherman 2013; Jacobs et al. 2009). The presence of leakier homes and dependence on natural ventilation can result in increased exposure to wildfire-related air pollutants through infiltration due to higher air exchange rates. These findings are consistent with those reported by (Wen and Burke 2022).

Several studies have conducted detailed association analyses between wildfire exposures and public health outcomes (e.g., see Reid et al. (2016)) and some have recently aimed to estimate causal effects of wildfire exposures on cognitive outcomes (Wen and Burke 2022; Cleland et al. 2022; Miller and Hui 2022; Gibbs et al. 2019). The analyses closest to ours is that of Wen and Burke (2022) which aimed to estimate the effect of exposure to PM_2.5_ during wildfires on academic performance in the United States. Wen and Burke (2022) reported that exposure to PM_2.5_ during wildfires led to a reduction in test scores of approximately 0.15% of a standard deviation, with more pronounced effects on younger students and across various socioeconomic and racial/ethnic groups. Since our analyses estimate the total effect of wildfire exposure (rather than solely PM_2.5_ derived from wildfires) on academic performance, our results are not directly comparable to those of Wen and Burke (2022). One advantage of estimating the total effect of wildfire exposure is that it offers a more natural causal interpretation. That is, we estimate the effect of an intervention that reduces one’s exposure wildfires rather than the effect of an intervention that reduces one’s exposure to PM_2.5_ (but not other pollutants) from wildfires. More generally, our use of the spatiotemporal causal inference framework of Christiansen et al. (2022) and detailed analyses on the spatiotemporal variability of the causal effects sets our analyses crucially apart from the rest of the literature in this domain.

It is also worthwhile discussing the challenges we faced in applying the causal inference framework of Christiansen et al. (2022). When using this framework, there is inherently an interplay between estimation accuracy and the assumptions required to causally interpret the results. That is, an analysis combining data across all spatial locations and at all time points offers more accurate estimation. However, this framework assumes that the average causal effect does not vary over time and space, which may be violated in an analysis combining all spatial locations and time points. Our subgroup analyses, which restricted the analyses to certain geographic regions and time points, explored this interplay between estimation accuracy and causal assumptions. Indeed, these analyses indicated a possible violation of the assumption of the causal effects not varying over time. However, the smaller sample sizes in these subgroup analyses necessitates that one interprets the results carefully. For this reason, we present both a global measure of the average causal effects as well separate average causal effect estimates in various subgroups.

Our study has a few limitations that should be considered. First, while the approach we applied to estimate the average causal effect of wildfire exposure on academic performance controls for all possible time-invariant confounders, there may exist time-varying confounders of the relationship between wildfire exposure and academic performance. Therefore, we must consider the possibility of some residual confounding bias. We could not adjust for time-varying confounders due to the limited data available in each municipality. Second, valid confidence intervals around the average causal effect estimates are not currently available, as the statistical literature has currently only developed methods to obtain p-values to test the null hypothesis of no average causal effect for this method. Third, considering that our study population consists of high school students aged between 15 and 18 years old, we do not have information on the students’ exposure to wildfire in the 15 years leading up to the test. This prior exposure could have affected their cognitive performance, which we are not able to account for in our analysis. Furthermore, we have not taken into account the potential effects of home exposures and infrastructure conditions at schools, which can influence the level of exposure that students experience. Another limitation of our study relates to the fact that the ENEM is a non-compulsory national exam, which is used for admission to various universities in Brazil. As a result, the sample of students who choose to take the exam may be biased towards those who are more academically inclined or have specific aspirations for higher education. However, it is worth noting that over the years, the number of students taking the ENEM has increased significantly, as many universities now require it for admission. This suggests that the sample of students taking the exam may be becoming more representative of the broader population of high school students.

Our study has several strengths. First, this is the first study in Brazil linking wildfire exposure and academic performance. Second, we estimated these effects for five different academic subjects and in different geographical regions (by averages wildfire exposure levels and average income levels) and time periods. Third, this is first study in the international literature on wildfire exposure and academic performance using a formal causal inference framework for spatiotemporal data, allowing us to precisely define the causal effects we aim to estimate. Fourth, our analyses make considerably weaker assumptions on the causal structure of the data (e.g., we do not require knowledge of time-invariant confounders) and on the distribution of the data (e.g., we make no particular distributional assumptions) compared to those from prior works.

## 5. CONCLUSIONS

Our study has shown that the impact of wildfires negatively affects the academic performance in most subjects and disproportionally affects lower socioeconomic regions in Brazil, and the effects worsened over time. Further research is needed to identify the mechanisms underlying these relationships to strengthen the evidence base and inform targeted policy interventions to mitigate the adverse impact of wildfires on educational outcomes and social inequality.

## Data Availability

The data that support the findings of this study are available from the corresponding author upon reasonable request.

## ACKNOWLEDGEMENT

SM was supported by the National Science Foundation Graduate Research Fellowship Program under Grant No. DGE2140743. WR was supported by the Brazilian Agencies National Council for Scientific and Technological Development (CNPq) and by the Ministry of Science, Technology and Innovation (MCTI). WCL was supported by the Higher Education Sprout Project of National Taiwan University (NTU-111L881004).

## CODE AVAILABILITY

The code for performing the statistical analyses is publicly available on GitHub (https://github.com/stmcg/wildfire).

## REFERENCES

Adetona, O., T. E. Reinhardt, J. Domitrovich, G. Broyles, A. M. Adetona, M. T. Kleinman, R. D. Ottmar, and L. P. Naeher. 2016. ‘Review of the health effects of wildland fire smoke on wildland firefighters and the public’, Inhal Toxicol, 28: 95–139.

Aguilera, Rosana, Thomas Corringham, Alexander Gershunov, and Tarik Benmarhnia. 2021. ‘Wildfire smoke impacts respiratory health more than fine particles from other sources: observational evidence from Southern California’, Nature Communications, 12: 1493.

Andersen, Zorana J., Peter Wahlin, Ole Raaschou-Nielsen, Thomas Scheike, and Steffen Loft. 2007. ‘Ambient particle source apportionment and daily hospital admissions among children and elderly in Copenhagen’, Journal of Exposure Science & Environmental Epidemiology, 17: 625–36.

Balmes, J. R. 2018. ‘Where There‘s Wildfire, There‘s Smoke’, N Engl J Med, 378: 881–83.

Bell, M. L., A. Zanobetti, and F. Dominici. 2014. ‘Who is more affected by ozone pollution? A systematic review and meta-analysis’, Am J Epidemiol, 180: 15–28.

Brook, R. D., S. Rajagopalan, C. A. Pope, 3rd, J. R. Brook, A. Bhatnagar, A. V. Diez-Roux, F. Holguin, Y. Hong, R. V. Luepker, M. A. Mittleman, A. Peters, D. Siscovick, S. C. Smith, Jr., L. Whitsel, J. D. Kaufman, Epidemiology American Heart Association Council on, Council on the Kidney in Cardiovascular Disease Prevention, Physical Activity Council on Nutrition, and Metabolism. 2010. ‘Particulate matter air pollution and cardiovascular disease: An update to the scientific statement from the American Heart Association’, Circulation, 121: 2331–78.

Calderon-Garciduenas, L., A. Mora-Tiscareno, E. Ontiveros, G. Gomez-Garza, G. Barragan-Mejia, J. Broadway, S. Chapman, G. Valencia-Salazar, V. Jewells, R. R. Maronpot, C. Henriquez-Roldan, B. Perez-Guille, R. Torres-Jardon, L. Herrit, D. Brooks, N. Osnaya-Brizuela, M. E. Monroy, A. Gonzalez-Maciel, R. Reynoso-Robles, R. Villarreal-Calderon, A. C. Solt, and R. W. Engle. 2008. ‘Air pollution, cognitive deficits and brain abnormalities: a pilot study with children and dogs’, Brain Cogn, 68: 117–27.

Chan, Wanyu R., Jeffrey Joh, and Max H. Sherman. 2013. ‘Analysis of air leakage measurements of US houses’, Energy and Buildings, 66: 616–25.

Christiansen, Rune, Matthias Baumann, Tobias Kuemmerle, Miguel D. Mahecha, and Jonas Peters. 2022. ‘Toward Causal Inference for Spatio-Temporal Data: Conflict and Forest Loss in Colombia’, Journal of the American Statistical Association, 117: 591–601.

Cleland, S. E., L. H. Wyatt, L. Wei, N. Paul, M. L. Serre, J. J. West, S. B. Henderson, and A. G. Rappold. 2022. ‘Short-Term Exposure to Wildfire Smoke and PM2.5 and Cognitive Performance in a Brain-Training Game: A Longitudinal Study of U.S. Adults’, Environ Health Perspect, 130: 67005.

Du, Xuan, Ye Wu, Lixin Fu, Shuxiao Wang, Shaojun Zhang, and Jiming Hao. 2012. ‘Intake fraction of PM2.5 and NOX from vehicle emissions in Beijing based on personal exposure data’, Atmospheric Environment, 57: 233–43.

Fan, J., S. Li, C. Fan, Z. Bai, and K. Yang. 2016. ‘The impact of PM2.5 on asthma emergency department visits: a systematic review and meta-analysis’, Environ Sci Pollut Res Int, 23: 843–50.

Forns, Joan, Payam Dadvand, Mikel Esnaola, Mar Alvarez-Pedrerol, Mònica López-Vicente, Raquel Garcia-Esteban, Marta Cirach, Xavier Basagaña, Mònica Guxens, and Jordi Sunyer. 2017. ‘Longitudinal association between air pollution exposure at school and cognitive development in school children over a period of 3.5 years’, Environmental Research, 159: 416–21.

Gibbs, Lisa, Jane Nursey, Janette Cook, Greg Ireton, Nathan Alkemade, Michelle Roberts, H. Colin Gallagher, Richard Bryant, Karen Block, Robyn Molyneaux, and David Forbes. 2019. ‘Delayed Disaster Impacts on Academic Performance of Primary School Children’, Child Development, 90: 1402–12.

Granger, C. W. J. 1980. ‘Testing for causality: A personal viewpoint’, Journal of Economic Dynamics and Control, 2: 329–52.

Guo, H., L. Morawska, C. He, Y. L. Zhang, G. Ayoko, and M. Cao. 2010. ‘Characterization of particle number concentrations and PM2.5 in a school: influence of outdoor air pollution on indoor air’, Environ Sci Pollut Res Int, 17: 1268–78.

Hernán, Miguel A, and James M Robins. 2010. “Causal inference.” In.: CRC Boca Raton, FL.

Jacobs, D. E., J. Wilson, S. L. Dixon, J. Smith, and A. Evens. 2009. ‘The relationship of housing and population health: a 30-year retrospective analysis’, Environ Health Perspect, 117: 597–604.

Laumbach, R. J., and H. M. Kipen. 2012. ‘Respiratory health effects of air pollution: update on biomass smoke and traffic pollution’, J Allergy Clin Immunol, 129: 3–11; quiz 12-3.

Lee, M., P. Koutrakis, B. Coull, I. Kloog, and J. Schwartz. 2016. ‘Acute effect of fine particulate matter on mortality in three Southeastern states from 2007-2011’, J Expo Sci Environ Epidemiol, 26: 173–9.

Liu, J. C., A. Wilson, L. J. Mickley, F. Dominici, K. Ebisu, Y. Wang, M. P. Sulprizio, R. D. Peng, X. Yue, J. Y. Son, G. B. Anderson, and M. L. Bell. 2017. ‘Wildfire-specific Fine Particulate Matter and Risk of Hospital Admissions in Urban and Rural Counties’, Epidemiology, 28: 77–85.

Martinez-Lazcano, J. C., E. Gonzalez-Guevara, M. del Carmen Rubio, J. Franco-Perez, V. Custodio, M. Hernandez-Ceron, C. Livera, and C. Paz. 2013. ‘The effects of ozone exposure and associated injury mechanisms on the central nervous system’, Rev Neurosci, 24: 337–52.

Miller, Rebecca K., and Iris Hui. 2022. ‘Impact of short school closures (1–5 days) on overall academic performance of schools in California’, Scientific Reports, 12: 2079.

Mohai, Paul, Byoung-Suk Kweon, Sangyun Lee, and Kerry Ard. 2011. ‘Air Pollution Around Schools Is Linked To Poorer Student Health And Academic Performance’, Health Affairs, 30: 852–62.

Pope, C. Arden, Michelle C. Turner, Richard T. Burnett, Michael Jerrett, Susan M. Gapstur, W. Ryan Diver, Daniel Krewski, and Robert D. Brook. 2015. ‘Relationships Between Fine Particulate Air Pollution, Cardiometabolic Disorders, and Cardiovascular Mortality’, Circulation Research, 116: 108–15.

Reid, C. E., M. Brauer, F. H. Johnston, M. Jerrett, J. R. Balmes, and C. T. Elliott. 2016. ‘Critical Review of Health Impacts of Wildfire Smoke Exposure’, Environ Health Perspect, 124: 1334–43.

Richmond-Bryant, J., C. Saganich, L. Bukiewicz, and R. Kalin. 2009. ‘Associations of PM2.5 and black carbon concentrations with traffic, idling, background pollution, and meteorology during school dismissals’, Sci Total Environ, 407: 3357–64.

Rivas, I., M. Viana, T. Moreno, M. Pandolfi, F. Amato, C. Reche, L. Bouso, M. Alvarez-Pedrerol, A. Alastuey, J. Sunyer, and X. Querol. 2014. ‘Child exposure to indoor and outdoor air pollutants in schools in Barcelona, Spain’, Environ Int, 69: 200–12.

Stenson, Chloe, Amanda J. Wheeler, Alison Carver, David Donaire-Gonzalez, Miguel Alvarado-Molina, Mark Nieuwenhuijsen, and Rachel Tham. 2021. ‘The impact of Traffic-Related air pollution on child and adolescent academic Performance: A systematic review’, Environment International, 155: 106696.

Suglia, S. Franco, A. Gryparis, R. O. Wright, J. Schwartz, and R. J. Wright. 2007. ‘Association of Black Carbon with Cognition among Children in a Prospective Birth Cohort Study’, American Journal of Epidemiology, 167: 280–86.

Sullivan, Andrew, Elaine Baker, and Tiina Kurvits. 2022. ‘Spreading like wildfire: The rising threat of extraordinary landscape fires‘.

Svendsen, E. R., M. Gonzales, S. Mukerjee, L. Smith, M. Ross, D. Walsh, S. Rhoney, G. Andrews, H. Ozkaynak, and L. M. Neas. 2012. ‘GIS-modeled indicators of traffic-related air pollutants and adverse pulmonary health among children in El Paso, Texas’, Am J Epidemiol, 176 Suppl 7: S131–41.

van Kempen, Elise, Paul Fischer, Nicole Janssen, Danny Houthuijs, Irene van Kamp, Stephen Stansfeld, and Flemming Cassee. 2012. ‘Neurobehavioral effects of exposure to traffic-related air pollution and transportation noise in primary schoolchildren’, Environmental Research, 115: 18–25.

Verma, Vishal, Andrea Polidori, James J. Schauer, Martin M. Shafer, Flemming R. Cassee, and Constantinos Sioutas. 2009. ‘Physicochemical and Toxicological Profiles of Particulate Matter in Los Angeles during the October 2007 Southern California Wildfires’, Environmental Science & Technology, 43: 954–60.

Weichenthal, S., M. Hatzopoulou, and M. S. Goldberg. 2014. ‘Exposure to traffic-related air pollution during physical activity and acute changes in blood pressure, autonomic and micro-vascular function in women: a cross-over study’, Part Fibre Toxicol, 11: 70.

Wen, Jeff, and Marshall Burke. 2022. ‘Lower test scores from wildfire smoke exposure’, Nature Sustainability, 5: 947–55.

Wiener, Norbert. 1956. ‘The theory of prediction’, Modern mathematics for engineers.

Wu, W., Y. Jin, and C. Carlsten. 2018. ‘Inflammatory health effects of indoor and outdoor particulate matter’, J Allergy Clin Immunol, 141: 833–44.

Xu, Q., X. Li, S. Wang, C. Wang, F. Huang, Q. Gao, L. Wu, L. Tao, J. Guo, W. Wang, and X. Guo. 2016. ‘Fine Particulate Air Pollution and Hospital Emergency Room Visits for Respiratory Disease in Urban Areas in Beijing, China, in 2013’, PLoS One, 11: e0153099.

Youssouf, H., C. Liousse, L. Roblou, E. M. Assamoi, R. O. Salonen, C. Maesano, S. Banerjee, and I. Annesi-Maesano. 2014. ‘Quantifying wildfires exposure for investigating healthrelated effects’, Atmospheric Environment, 97: 239–51.

