## Supplementary Tables 1-5 for "Wildfire exposure and academic performance in Brazil: a causal inference approach for spatiotemporal data"

| | |
|---|---|
| Supplemental Table 1 | Results of the sensitivity analysis restricting the modelled mean SAP values in the municipality-specific regression models to the range 0 to 800. |
| Supplemental Table 2 | Results of the sensitivity analysis restricting the modelled mean SAP values in the municipality-specific regression models to the range 200 to 1000. |
| Supplemental Table 3 | Results of the sensitivity analysis restricting the modelled mean SAP values in the municipality-specific regression models to the range 200 to 800. |
| Supplemental Table 4 | Results of the sensitivity analysis that perform a log transformation of the wildfire density in the municipality-specific regression models. |
| Supplemental Table 5 | Results of the sensitivity analysis which estimates the average causal effect by taking the sample median (rather than the sample mean) of the municipality-specific estimates. |

**Supplemental Table 1**. Results of the sensitivity analysis restricting the modelled mean SAP values in the municipality-specific regression models to the range 0 to 800.

| | ACE Estimate (p-value) | | | | |
|---|---|---|---|---|---|
| **Subgroup** | **Natural Sciences** | **Human Sciences** | **Language** | **Mathematics** | **Essay** |
| All municipalities | -5.82 (0.02) | -11.63 (0.01) | -4.46 (0.15) | -8.71 (0.07) | 6.00 (0.01) |
| By year (2010) | | | | | |
| Pre 2010[1] | -0.69 (0.66) | 9.98 (0.02) | 3.18 (0.20) | 6.17 (0.03) | 4.69 (0.07) |
| Post 2010 | -5.17 (0.30) | -19.13 (0.01) | 1.40 (0.16) | -11.05 (0.14) | -16.41 (0.02) |
| Change | -4.49 (0.36) | -29.12 (0.01) | -1.77 (0.60) | -17.22 (0.03) | -21.10 (0.01) |
| By year (2011) | | | | | |
| Pre 2011 | -1.68 (1.00) | 18.18 (0.01) | 5.22 (0.10) | 2.33 (0.25) | 9.50 (0.01) |
| Post 2011 | -4.31 (0.65) | -7.96 (0.40) | -7.15 (0.24) | -21.09 (0.02) | -20.78 (0.01) |
| Change | -2.63 (0.84) | -26.15 (0.01) | -12.37 (0.02) | -23.42 (0.01) | -30.28 (0.01) |
| By year (2012) | | | | | |
| Pre 2012 | -2.04 (0.96) | 6.03 (0.03) | 4.05 (0.10) | -4.31 (0.85) | 8.65 (0.01) |
| Post 2012 | -16.36 (0.02) | -22.32 (0.02) | -16.79 (0.02) | -25.51 (0.01) | -25.77 (0.01) |
| Change | -14.31 (0.04) | -28.34 (0.01) | -20.83 (0.01) | -21.20 (0.02) | -34.42 (0.01) |
| By year (2013) | | | | | |
| Pre 2013 | 1.17 (0.42) | 2.51 (0.21) | 6.36 (0.01) | -8.76 (0.11) | 15.00 (0.01) |
| Post 2013 | -5.85 (0.81) | -18.68 (0.02) | -6.55 (0.75) | -9.96 (0.67) | -19.45 (0.19) |
| Change | -7.02 (0.65) | -21.19 (0.01) | -12.91 (0.14) | -1.20 (0.78) | -34.45 (0.01) |
| By average exposure level[2] | | | | | |
| Below the median | -8.71 (0.28) | -20.56 (0.03) | -9.55 (0.11) | -14.48 (0.16) | -0.42 (0.03) |
| Above the median | -3.09 (0.07) | -3.20 (0.10) | 0.35 (0.99) | -3.27 (0.13) | 12.07 (0.01) |
| Change | 5.61 (0.77) | 17.36 (0.10) | 9.90 (0.15) | 11.21 (0.64) | 12.49 (0.64) |
| By average income level[3] | | | | | |
| Below the median | -6.09 (0.08) | -11.14 (0.01) | -5.98 (0.10) | -11.71 (0.01) | 5.20 (0.02) |
| Above the median | -5.69 (0.56) | -14.79 (0.03) | -0.15 (0.59) | 1.37 (0.33) | 8.04 (0.01) |
| Change | 0.40 (0.72) | -3.65 (0.72) | 5.84 (0.18) | 13.08 (0.04) | 2.85 (0.62) |

[1] The subgroup "Pre 2010" includes all data before and including 2010. The subgroups "Pre 2011", "Pre 2012", and "Pre 2013" were defined in the same manner.
[2] The median wildfire density was 0.0125 wildfires/$km^2$.
[3] The median income level was R$797.71 (currency of Brazil).

**Supplemental Table 2**. Results of the sensitivity analysis restricting the modelled mean SAP values in the municipality-specific regression models to the range 200 to 1000.

| | ACE Estimate (p-value) | | | | |
|---|---|---|---|---|---|
| **Subgroup** | **Natural Sciences** | **Human Sciences** | **Language** | **Mathematics** | **Essay** |
| All municipalities | -1.17 (0.26) | -4.88 (0.02) | -0.66 (0.38) | -1.05 (0.25) | 19.50 (0.01) |
| By year (2010) | | | | | |
| Pre 2010[1] | 5.84 (0.48) | 18.87 (0.01) | 10.10 (0.05) | 15.28 (0.02) | 12.74 (0.03) |
| Post 2010 | 1.82 (0.75) | -7.57 (0.02) | 7.12 (0.06) | 2.97 (0.85) | -1.05 (0.38) |
| Change | -4.02 (0.49) | -26.44 (0.01) | -2.98 (0.49) | -12.31 (0.06) | -13.79 (0.02) |
| By year (2011) | | | | | |
| Pre 2011 | 3.40 (0.67) | 26.87 (0.01) | 9.64 (0.06) | 9.38 (0.11) | 20.06 (0.01) |
| Post 2011 | 4.86 (0.67) | 3.30 (0.75) | 1.89 (0.84) | -5.17 (0.06) | -3.23 (0.11) |
| Change | 1.45 (1.00) | -23.57 (0.01) | -7.75 (0.06) | -14.54 (0.02) | -23.29 (0.01) |
| By year (2012) | | | | | |
| Pre 2012 | 3.31 (0.71) | 13.40 (0.02) | 7.97 (0.04) | 2.86 (0.82) | 21.44 (0.01) |
| Post 2012 | -4.83 (0.06) | -8.97 (0.03) | -6.80 (0.04) | -8.20 (0.02) | -7.47 (0.11) |
| Change | -8.14 (0.08) | -22.37 (0.01) | -14.76 (0.02) | -11.06 (0.06) | -28.91 (0.01) |
| By year (2013) | | | | | |
| Pre 2013 | 6.15 (0.19) | 7.88 (0.08) | 10.28 (0.01) | -1.82 (0.26) | 29.41 (0.01) |
| Post 2013 | 7.85 (0.61) | -6.98 (0.11) | 5.53 (0.64) | 7.43 (0.79) | -2.51 (0.57) |
| Change | 1.70 (0.96) | -14.85 (0.02) | -4.75 (0.31) | 9.25 (0.48) | -31.92 (0.01) |
| By average exposure level[2] | | | | | |
| Below the median | 1.67 (0.77) | -4.05 (0.12) | -0.70 (0.37) | 3.43 (0.88) | 33.54 (0.01) |
| Above the median | -3.85 (0.05) | -5.66 (0.03) | -0.62 (0.83) | -5.28 (0.04) | 6.24 (0.02) |
| Change | -5.52 (0.65) | -1.61 (0.68) | 0.09 (0.49) | -8.71 (0.51) | -27.30 (0.02) |
| By average income level[3] | | | | | |
| Below the median | -1.94 (0.18) | -4.09 (0.07) | -2.15 (0.21) | -4.30 (0.06) | 19.61 (0.01) |
| Above the median | 0.72 (0.81) | -9.01 (0.05) | 3.54 (0.54) | 9.88 (0.18) | 18.18 (0.01) |
| Change | 2.66 (0.65) | -4.92 (0.52) | 5.69 (0.29) | 14.19 (0.04) | -1.42 (0.96) |

[1] The subgroup "Pre 2010" includes all data before and including 2010. The subgroups "Pre 2011", "Pre 2012", and "Pre 2013" were defined in the same manner.

[2] The median wildfire density was 0.0125 wildfires/$km^2$.

[3] The median income level was R$797.71 (currency of Brazil).

**Supplemental Table 3**. Results of the sensitivity analysis restricting the modelled mean SAP values in the municipality-specific regression models to the range 200 to 800.

| | ACE Estimate (p-value) | | | | |
|---|---|---|---|---|---|
| **Subgroup** | **Natural Sciences** | **Human Sciences** | **Language** | **Mathematics** | **Essay** |
| All municipalities | -2.76 (0.19) | -7.08 (0.01) | -1.81 (0.39) | -3.60 (0.25) | 11.45 (0.01) |
| By year (2010) | | | | | |
| Pre 2010[1] | 1.91 (0.61) | 14.03 (0.01) | 6.75 (0.05) | 9.34 (0.02) | 7.00 (0.04) |
| Post 2010 | -0.63 (0.64) | -12.22 (0.01) | 4.54 (0.06) | -2.83 (0.57) | -6.81 (0.29) |
| Change | -2.55 (0.57) | -26.24 (0.01) | -2.21 (0.53) | -12.17 (0.06) | -13.81 (0.02) |
| By year (2011) | | | | | |
| Pre 2011 | 1.00 (0.80) | 20.35 (0.01) | 6.62 (0.07) | 5.13 (0.14) | 12.29 (0.01) |
| Post 2011 | 1.07 (0.78) | -2.61 (0.79) | -2.04 (0.61) | -10.75 (0.04) | -9.09 (0.11) |
| Change | 0.07 (1.00) | -22.97 (0.01) | -8.66 (0.06) | -15.88 (0.02) | -21.38 (0.01) |
| By year (2012) | | | | | |
| Pre 2012 | -0.10 (0.97) | 8.48 (0.02) | 5.44 (0.06) | -0.63 (0.92) | 12.65 (0.01) |
| Post 2012 | -8.86 (0.03) | -14.20 (0.02) | -11.38 (0.03) | -13.82 (0.02) | -13.85 (0.06) |
| Change | -8.75 (0.10) | -22.68 (0.01) | -16.83 (0.01) | -13.19 (0.06) | -26.50 (0.01) |
| By year (2013) | | | | | |
| Pre 2013 | 3.05 (0.29) | 4.38 (0.15) | 7.71 (0.01) | -5.22 (0.17) | 19.69 (0.01) |
| Post 2013 | 0.76 (0.85) | -11.22 (0.06) | -0.38 (0.93) | -1.67 (0.88) | -7.62 (0.53) |
| Change | -2.29 (0.86) | -15.60 (0.02) | -8.09 (0.21) | 3.55 (0.64) | -27.31 (0.01) |
| By average exposure level[2] | | | | | |
| Below the median | -1.76 (0.71) | -11.08 (0.06) | -3.83 (0.28) | -3.93 (0.61) | 14.49 (0.01) |
| Above the median | -3.71 (0.05) | -3.30 (0.10) | 0.09 (1.00) | -3.29 (0.14) | 8.58 (0.01) |
| Change | -1.95 (0.66) | 7.77 (0.33) | 3.92 (0.38) | 0.64 (0.79) | -5.90 (0.08) |
| By average income level[3] | | | | | |
| Below the median | -3.35 (0.14) | -6.47 (0.04) | -3.20 (0.19) | -6.25 (0.04) | 11.21 (0.01) |
| Above the median | -1.54 (0.84) | -10.58 (0.06) | 2.02 (0.49) | 5.17 (0.18) | 11.44 (0.01) |
| Change | 1.81 (0.63) | -4.11 (0.64) | 5.22 (0.22) | 11.41 (0.06) | 0.23 (0.76) |

[1] The subgroup "Pre 2010" includes all data before and including 2010. The subgroups "Pre 2011", "Pre 2012", and "Pre 2013" were defined in the same manner.

[2] The median wildfire density was 0.0125 wildfires/km$^2$.

[3] The median income level was R$797.71 (currency of Brazil).

**Supplemental Table 4**. Results of the sensitivity analysis that perform a log transformation of the wildfire density in the municipality-specific regression models.

| | ACE Estimate (p-value) | | | | |
|---|---|---|---|---|---|
| **Subgroup** | **Natural Sciences** | **Human Sciences** | **Language** | **Mathematics** | **Essay** |
| All municipalities | -5.62 (0.01) | -6.44 (0.01) | -1.74 (0.41) | -4.70 (0.10) | 14.91 (0.01) |
| By year (2010) | | | | | |
| Pre 2010[1] | 0.49 (0.89) | 21.73 (0.01) | 7.14 (0.15) | 15.25 (0.02) | 17.78 (0.01) |
| Post 2010 | -0.75 (0.77) | -7.61 (0.02) | 4.54 (0.04) | -5.88 (0.12) | -8.14 (0.13) |
| Change | -1.24 (0.78) | -29.34 (0.01) | -2.60 (0.53) | -21.12 (0.02) | -25.91 (0.01) |
| By year (2011) | | | | | |
| Pre 2011 | -4.57 (0.29) | 27.63 (0.01) | 4.91 (0.23) | 4.77 (0.40) | 20.79 (0.01) |
| Post 2011 | -1.20 (0.72) | 1.85 (0.61) | -0.01 (1.00) | -13.47 (0.01) | -10.13 (0.06) |
| Change | 3.37 (0.54) | -25.78 (0.01) | -4.92 (0.31) | -18.25 (0.01) | -30.92 (0.01) |
| By year (2012) | | | | | |
| Pre 2012 | -2.20 (0.55) | 13.66 (0.01) | 4.86 (0.14) | -0.27 (0.87) | 20.99 (0.01) |
| Post 2012 | -9.66 (0.05) | -9.72 (0.05) | -9.56 (0.03) | -20.12 (0.01) | -16.60 (0.02) |
| Change | -7.46 (0.17) | -23.38 (0.01) | -14.42 (0.02) | -19.85 (0.02) | -37.59 (0.01) |
| By year (2013) | | | | | |
| Pre 2013 | 0.42 (0.81) | 6.82 (0.05) | 8.89 (0.01) | -4.37 (0.18) | 27.26 (0.01) |
| Post 2013 | -0.32 (1.00) | -8.50 (0.17) | -1.63 (0.84) | -1.32 (1.00) | -9.98 (0.33) |
| Change | -0.74 (0.98) | -15.31 (0.03) | -10.52 (0.10) | 3.06 (0.63) | -37.24 (0.01) |
| By average exposure level[2] | | | | | |
| Below the median | -3.13 (0.36) | -7.99 (0.04) | -2.60 (0.29) | -2.30 (0.69) | 8.78 (0.09) |
| Above the median | -7.98 (0.01) | -4.98 (0.15) | -0.93 (0.74) | -6.98 (0.04) | 20.71 (0.01) |
| Change | -4.85 (0.19) | 3.01 (0.73) | 1.67 (0.74) | -4.68 (0.33) | 11.93 (0.14) |
| By average income level[3] | | | | | |
| Below the median | -6.84 (0.01) | -7.71 (0.02) | -4.12 (0.12) | -10.28 (0.01) | 15.48 (0.01) |
| Above the median | -1.90 (0.60) | -3.05 (0.37) | 5.88 (0.06) | 14.66 (0.01) | 12.54 (0.02) |
| Change | 4.94 (0.24) | 4.66 (0.34) | 9.99 (0.01) | 24.95 (0.01) | -2.93 (0.75) |

[1] The subgroup “Pre 2010” includes all data before and including 2010. The subgroups “Pre 2011”, “Pre 2012”, and “Pre 2013” were defined in the same manner.

[2] The median wildfire density was 0.0125 wildfires/$km^2$.

[3] The median income level was R$797.71 (currency of Brazil).

**Supplemental Table 5**. Results of the sensitivity analysis which estimates the average causal effect by taking the sample median (rather than the sample mean) of the municipality-specific estimates.

| | ACE Estimate (p-value) | | | | |
|---|---|---|---|---|---|
| **Subgroup** | **Natural Sciences** | **Human Sciences** | **Language** | **Mathematics** | **Essay** |
| All municipalities | -1.76 (0.84) | -1.99 (0.65) | -0.24 (0.45) | -4.36 (0.29) | 9.14 (0.01) |
| By year (2010) | | | | | |
| Pre 2010[1] | -2.53 (0.30) | 13.96 (0.01) | 4.25 (0.05) | 6.12 (0.02) | 4.44 (0.07) |
| Post 2010 | 1.08 (0.25) | -3.56 (0.10) | 3.06 (0.02) | 1.57 (0.16) | 0.04 (0.96) |
| Change | 3.60 (0.15) | -17.52 (0.01) | -1.19 (0.79) | -4.55 (0.63) | -4.40 (0.25) |
| By year (2011) | | | | | |
| Pre 2011 | -0.48 (0.92) | 16.42 (0.01) | 4.83 (0.02) | 4.77 (0.08) | 8.00 (0.01) |
| Post 2011 | 2.22 (0.10) | 1.28 (0.29) | -1.10 (0.96) | -6.14 (0.18) | -5.54 (0.16) |
| Change | 2.71 (0.22) | -15.14 (0.01) | -5.93 (0.10) | -10.92 (0.03) | -13.53 (0.01) |
| By year (2012) | | | | | |
| Pre 2012 | -0.74 (0.90) | 10.12 (0.01) | 4.72 (0.02) | 2.47 (0.22) | 12.87 (0.01) |
| Post 2012 | -1.77 (0.43) | -3.34 (0.23) | -3.51 (0.14) | -4.53 (0.29) | -9.85 (0.10) |
| Change | -1.03 (0.57) | -13.46 (0.01) | -8.24 (0.01) | -7.00 (0.07) | -22.72 (0.01) |
| By year (2013) | | | | | |
| Pre 2013 | -0.54 (0.62) | 4.84 (0.01) | 4.18 (0.01) | -3.58 (0.30) | 11.74 (0.01) |
| Post 2013 | 4.89 (0.08) | -2.75 (0.31) | 0.89 (0.52) | 2.18 (0.71) | -1.53 (0.73) |
| Change | 5.44 (0.30) | -7.59 (0.02) | -3.30 (0.20) | 5.76 (0.41) | -13.26 (0.02) |
| By average exposure level[2] | | | | | |
| Below the median | -5.49 (0.37) | -2.89 (0.45) | 0.57 (0.21) | -5.30 (0.95) | 25.21 (0.01) |
| Above the median | -0.11 (0.65) | 1.45 (0.18) | 0.95 (0.52) | -3.43 (0.07) | 6.89 (0.01) |
| Change | 5.38 (0.64) | 4.34 (0.17) | 0.38 (0.39) | 1.87 (0.33) | -18.32 (0.01) |
| By average income level[3] | | | | | |
| Below the median | -1.66 (0.81) | -1.85 (0.62) | -1.50 (0.92) | -4.84 (0.16) | 11.32 (0.01) |
| Above the median | 4.40 (0.10) | -0.95 (0.55) | 4.50 (0.06) | 12.62 (0.01) | 12.05 (0.01) |
| Change | 6.05 (0.14) | 0.90 (0.82) | 6.00 (0.15) | 17.45 (0.01) | 0.74 (0.46) |

[1] The subgroup "Pre 2010" includes all data before and including 2010. The subgroups "Pre 2011", "Pre 2012", and "Pre 2013" were defined in the same manner.

[2] The median wildfire density was 0.0125 wildfires/km$^2$.

[3] The median income level was R$797.71 (currency of Brazil).
